# Prevalence of various inappropriate antibiotic doses among pediatric patients of in-patient, out-patient, and emergency care units in Bangladesh: a cross-sectional study

**DOI:** 10.1101/2022.06.21.22276716

**Authors:** A. F. M. Mahmudul Islam, Md Abu Raihan, Md. Galib Ishraq Emran, Khandaker Tanveer Ahmed, Nahria Amin Nusrat, Md. Asif Hasan, Anika Bushra Lamisa, Tania Ahmed, Halima Akter Happy, Mahfuza Khatoon

## Abstract

Antibiotics are the most frequently recommended medications for the treatment of bacterial infections. In most instances, pediatric patients have been prescribed antibiotics without a suitable dosage. Furthermore, existing antibiotic dosages in the market are not appropriate for pediatric patients. As a result, the complication brought on by antibiotic resistance increases day by day. This study was conducted to examine and evaluate the appropriateness of antibiotic doses. This study used a convenient sampling method to collect 300 filled prescription orders from heterogeneous pediatric patients prescribed by physicians from three different patient care departments: emergency patient care, inpatient care, and outpatient care, of various clinics, hospitals, and health care centers. There were 165 male responders and 135 female respondents in all. This study rated the 12 most prevalent diseases diagnosed in pediatric patients of various ages. This study revealed that prescribers recommended antibiotic overdoses for patients in the outdoor unit and that antibiotic underdoses were more prevalent among pediatric patients in the outdoor unit. According to this finding, it is straightforward to conclude that prescribers underestimated the doses and could not change the dosage for kids of early childhood and toddler age. Unfamiliar prescribers with the correct pediatric dose standards may have committed prescription dosing errors repeatedly, leading to the development of multi-drug resistance. More research can be conducted on these antibiotics’ effects or potential adverse effects on pediatric patients.

## Introduction

Antibiotics are the most promising medications prescribed for treating bacterial infections. Antibiotics are frequently prescribed to pediatric patients for various infectious diseases. The age groups of pediatric patients include neonatal, infancy, toddler, early childhood, middle childhood, early adolescence, and late adolescence (1).

Most pediatric patients are admitted to the hospital to treat infectious diseases. Two-thirds of pediatric patients were brought to the hospital’s emergency unit due to contagious diseases (2, 3). Other studies demonstrated that 12-18% of hospitalized pediatric patients received antibiotics for emergency care (4, 5). In 30-50% of the cases, it was discovered that inappropriate antibiotics were prescribed (6, 7). Although appropriate antibiotics are prescribed in some instances, doses are frequently incorrect, either overdose or underdose. An error in antibiotic dosage has the same effect as the use of inappropriate antibiotics. Antimicrobial resistance, one of the world’s primary concerns, is caused by dosage errors and improper antibiotic use (8). In addition, this can result in drug-related side effects, changes in the body’s normal flora, sensitization to the possibility of future allergies, and an increased risk of asthma or obesity in children (9, 10).

During childhood, the development of organisms and the immune system makes drug dosing for children more complicated and problematic than for adults. Changes in pharmacokinetics (how drugs are absorbed, distributed, metabolized and eliminated by the body) affect pharmacodynamics (the actions of the drug on the body) (11). The optimal antibiotic dose is heavily dependent on the development of renal function and hepatic metabolism during the early stages of life. Any error in antibiotic dosing may result in numerous adverse effects, particularly diarrhea, and may increase bacterial resistance (12).

It is more challenging to formulate antibiotic dosages for children than for adults. Antibiotic doses must be formulated for achieving optimal effectiveness (13). The available antibiotic dosages in the market are designed for adults. To achieve the correct dosage for children, tablets may need to be split or crushed (14). Although liquid antibiotics are easier to measure, measuring small volumes with a cup or spoon can be inaccurate (15). Typically, syringes are more precise than cups or spoons for correct dosing (16), and they can be used to improve precision (17).

It is also difficult to formulate the proper antibiotic dosage because some antibiotics are unpalatable (18). As a result of malnutrition, most children in developing nations are underweight, and antibiotic doses are not always proportional to their body weight. Therefore, antibiotic dosages may be incorrect, even if they become ineffective after administration.

The correct antibiotic dosage for pediatric patients is crucial for preventing adverse effects and bacterial resistance development. Parents must be careful to use the proper antibiotics when splitting or crushing antibiotic tablets and measuring liquid antibiotics before administration. Before prescribing antibiotics to pediatric patients, physicians must be cautious about antibiotic dosage. For the safety and efficacy of antibiotics, pediatric patients must receive the correct antibiotic dosage.

### Rationality and objectives

The principles of rational antibiotic use are well-defined, and inappropriate prescribing patterns are reported for adults, but there is insufficient data regarding pediatric patients’ dose errors. Accurate information on antibiotics in pediatric patients is essential for enhancing the quality of antibiotic prescription practices, preventing antibiotic resistance, and avoiding side effects. This study was conducted to explore the incidence of serious errors of prescribing inappropriate doses of different antibiotics in pediatric patients. This study also investigated and examined whether the doses of antibiotics were appropriate or inappropriate for the pediatric patient according to his/her age and body weight against particular diseases.

## Materials and method

### Study design and setting

A cross-sectional study was conducted on pediatric patients in three different care units, including inpatient, outpatient, and emergency care units, who adhered to different antibiotic dosage regimens. During the study, we collected pediatric patients’ prescriptions. We consulted with their respective caregivers to collect information such as age category, gender, patient type, most frequently encountered diseases, daily frequency of antibiotic administration, antibiotic doses, antibiotic dose forms, and dose regimens for pediatric patients. The research was conducted in hospitals in the Savar subdistrict of Dhaka, Bangladesh.

### Inclusion and exclusion criteria while selecting pediatric patients

In this study, mild-to-moderate infections in pediatric patients were preserved. Patients who presented with severe infection or sepsis or who were immunocompromised were not chosen as candidates for evaluating the appropriateness of antibiotic dosing. In addition, patients with abnormal kidney or liver function were not considered for antibiotic dosing evaluation. After all of these conditions were met, the same number of pediatric patients from each of the three care units was chosen to see if the antibiotic doses were proper.

### Sampling method and recruiting process

In clinical research, non-probability sampling methods were utilized because there was no sampling frame of compatible patients for the study. The most common sampling method for clinical research is convenient sampling, in which participants are selected from the sampling population based on their accessibility and availability (19). This cross-sectional clinical survey collected data on pediatric patients and their prescribed antibiotics. This clinical study surveyed recruited participants, consisting of all pediatric categorical patients from all three care units in selected hospitals in the Savar subdistrict. This clinical study recruited only patients younger than 14 years old. We selected these individuals based on previously established inclusion criteria for pediatric patients accompanied by their caregivers or parents during treatment. Face-to-face interviews were used to conduct every aspect of the interview procedure. The entire study was conducted using printed versions of previously designed questionnaires with the parents or caregivers of pediatric patients receiving treatment. We only took on children as patients if their parents or other caregivers were involved in their treatment or already knew what was going on.

### Study area & sample size

The study was conducted in the Savar region, where sampled patients from three hospitals who met the criteria mentioned earlier and were willing to volunteer their prescription information. During the study period, 300 antibiotic-prescribed pediatric patients were selected as study samples from 714 pediatric patients treated in these three hospitals’ inpatient care units, outpatient care units, and emergency care units.

### Ethical statement

All procedures of the current study was carried out according to the principle of human investigation by the Institutional Research Ethics Committee’s ethical guidelines. Formal ethical approval was granted to the Ethical Clearance Committee, the ethical review board of Gono Bishwabidyalay, Savar, Dhaka, Bangladesh (Reference No. CMR/E/002). The objective and methodology of the study were communicated to all participants. Before enrollment, written and informed consent of parents and caregivers was obtained from all eligible participants. Ensuring privacy and confidentiality was given more significant consideration throughout the study by concealing the patient’s identity. Using the coding system, all data were gathered anonymously and analyzed.

### Study procedures

We conducted this study between 6 January 2021 and 28 May 2021 to evaluate the adequacy of antibiotic dosing. In addition, to check and assess the appropriateness of dosing for prescribed antibiotics, we must compare the antibiotic dosing for a specific age and weight of the pediatric population selected by the prescriber for a particular antibiotic with the dosing recommended by the most recent version of the Harriet Lane Handbook, a pediatric drug reference (19). We assumed an antibiotic overdose when the calculated weight-based daily dose (mg/kg/day) exceeded the Harriet Lane Handbook’s (20) maximum recommended pediatric dose for a specific antibiotic (20). When the prescribed weight-based daily dose (mg/kg/day) was less than the recommended pediatric dose in our reference book, we assumed the antibiotic was administered sub-therapeutically or insufficiently. If any antibiotic dosage was an overdose or underdose, we assumed that the antibiotic dosage in this study was inappropriate.

This study selected and included antibiotic prescriptions for pediatric patients younger than 14 years of age, indicating the study’s focus on the heterogenous pediatric population in Bangladesh. According to NICHD (National Institute of Child Health and Human Development) pediatric terminology, childhood is divided into five age ranges such as a preterm newborn infant, term neonatal (birth to 27days), infancy (28days to 12months), toddlers (13months to 2years), early childhood (2 to 5 years), middle childhood (6-11 years), and adolescent (12 to 18 years), etc. This population can also be referred to as heterogeneous. Approximately 165 of the total pediatric patients were male, while 135 were female.

### Questionnaire details

A questionnaire (See <u>S1 File</u>) was designed and developed to investigate and assess the appropriateness or inappropriateness of antibiotic dosing in pediatric patients. The questionnaire was translated into the Bangla format to prevent further errors and facilitate better comprehension and response (See <u>S2 File</u>). The questionnaire was divided into two sections. The primary focus of the first section is on the demographic profiles of pediatric patients. The second section of the study questionnaire focuses solely on the medication history to organize the list of antibiotics and the diseases for which they were prescribed. The questionnaire contained nine semi-closed questions to elicit more specific responses to the text and two multiple-choice questions to identify the care units and their gender.

### Statistical data analysis

All statistical analyses were conducted using Microsoft Excel 2019 and RStudio 1.3.959. The data was cleansed, edited, and arranged using Microsoft Excel. The data were then imported into RStudio via an Excel file. For all socio-demographic factors and antibiotics, with frequency percentages were displayed. Cross-tabulations were utilized to understand the relationship between dose level and frequency of administration and patient type, age group, dosage regimen, and dosage form. Then, chi-square tests were conducted to determine significance, and only significant dependencies were reported. To indicate the dose level that deviated from the standard level, median absolute deviations for various dose levels were computed, and Kruskal-Wallis tests were used to assess the significance of the deviations.

### Participant’s consent

As the subjects of this study were children, the majority of whom were 14 years of age or younger, it was deemed appropriate to accept the consent of the parents or guardians of pediatric patients as their authorized representative. An informed Consent Form (ICF) was provided to the parent or caretaker of the selected individual in both Bangla and English to ensure their comprehension. The informed Consent Form contains all pertinent information, including specific objectives, interview duration, selection criteria, signature option for the legal guardian of selected individuals, etc. Additionally, we prioritized and ensured the confidentiality of their information for this research study. If the subjects’ guardians are illiterate, our investigators read and explain the content of the Informed Consent Form to ensure their comprehension. Before beginning the formal interview, we obtained their handwritten signature after they had voluntarily consented to participate in this research without any coercion or inducement. All of the selected participants (parents or caregivers) volunteered all of their current medical and medication history for this study during the face-to-face interviewing process. During this study, the informed parents, caregivers, or attendees of participants were also willing to answer questions consisting of personal information, disease description, and prescription to collect data regarding antibiotic dosages.

## Result

### Patient’s demographic and disease information

The study was carried out among 300 pediatric patients. They were categorized on the basis of age, sex and type (outdoor, indoor & emergency patients). Majority of the patients were term neonatal (33.3%) and male (55%). Almost equal number of samples have been collected from indoor (33.7%), outdoor (33.3%) and emergency (33.0%) patients. Most of the patients were suffering from different kinds of fevers (22%), pneumonia (17%), cough (16%), delayed crying (14%) and cold (13.3%). Table 1 presents detailed information.

**Table 1.**
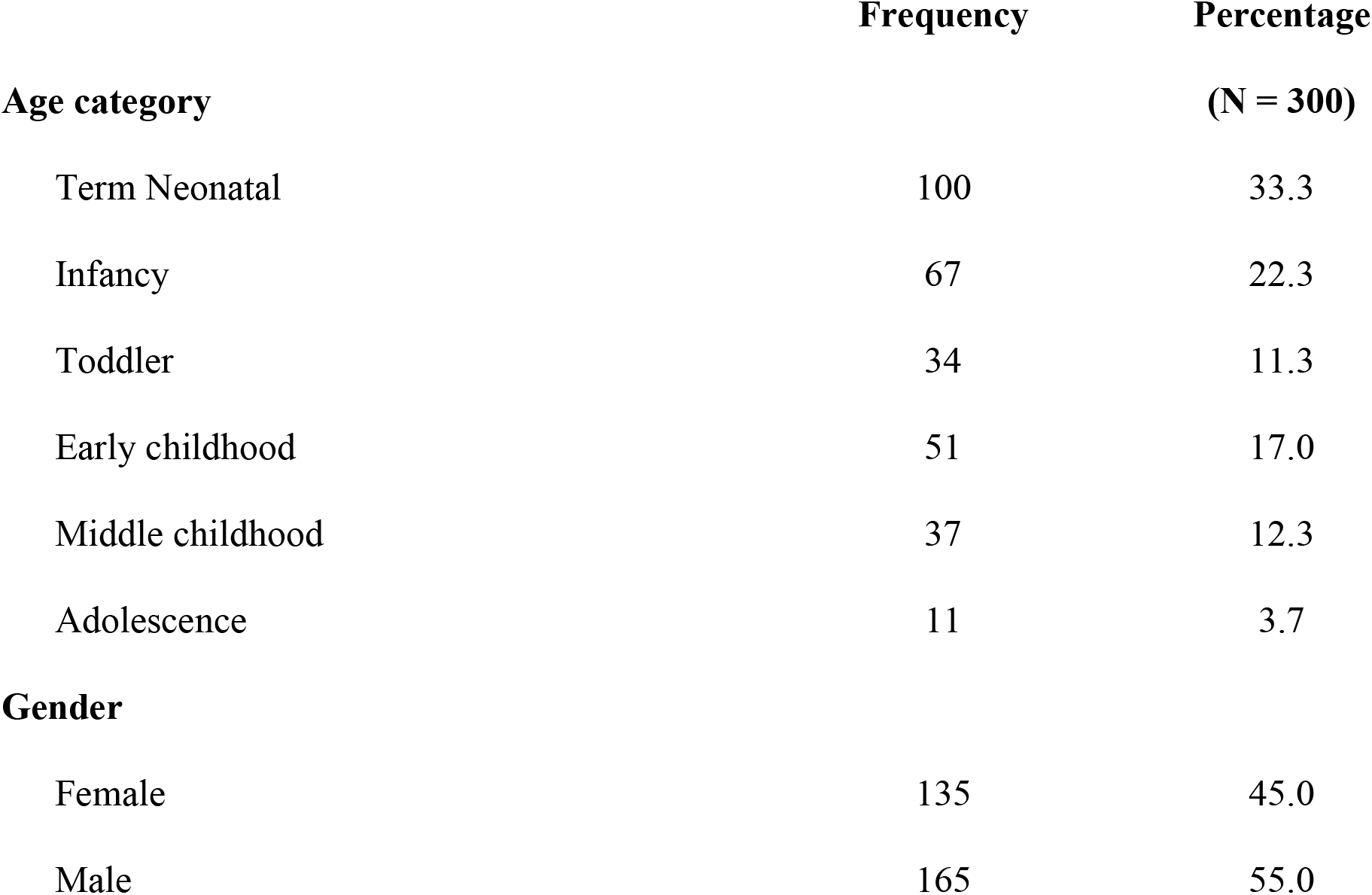
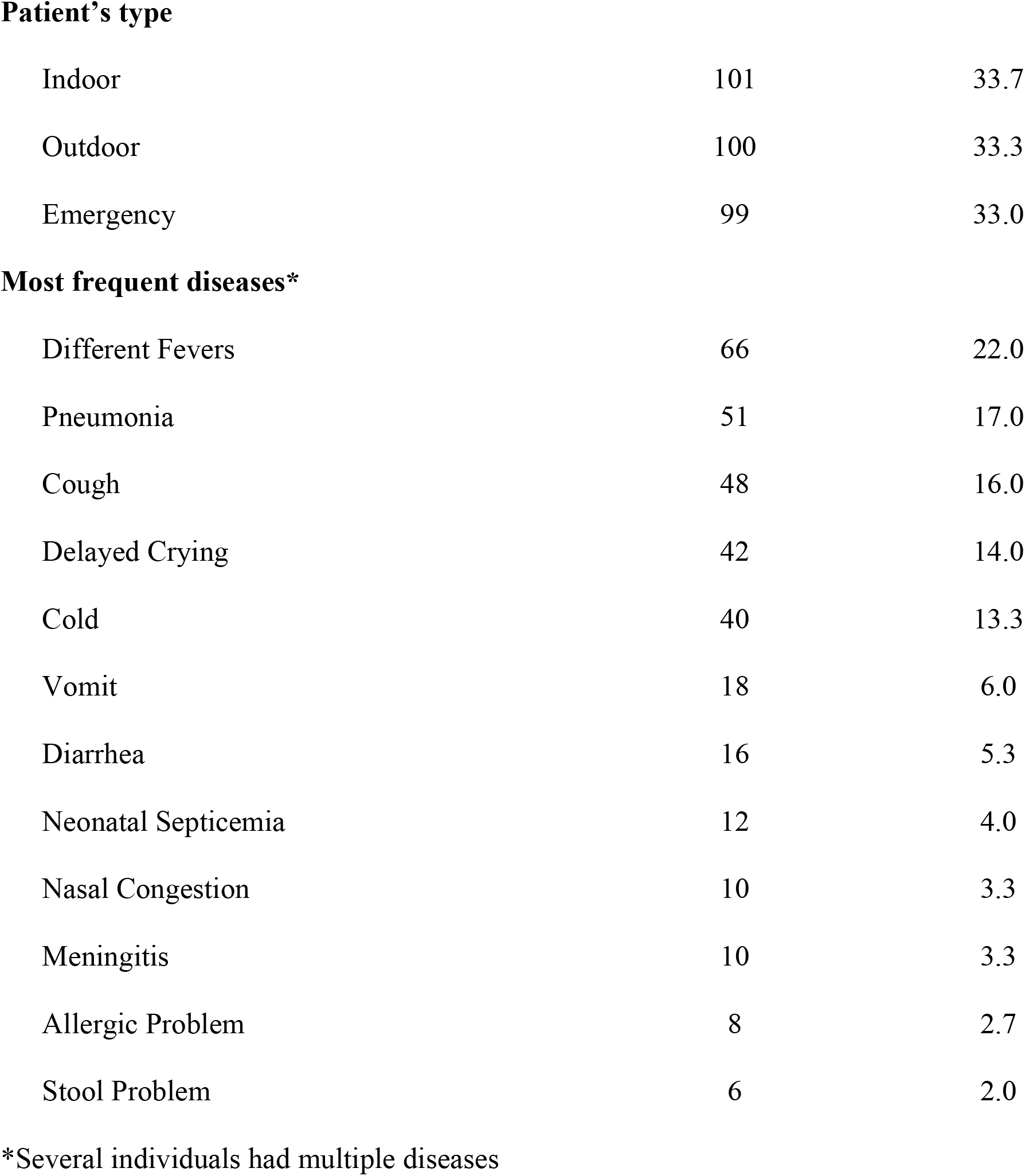
Participated patient’s demographic and disease information

|  | Frequency | Percentage |
| --- | --- | --- |
| Age category |  | (N = 300) |
| Term Neonatal | 100 | 33.3 |
| Infancy | 67 | 22.3 |
| Toddler | 34 | 11.3 |
| Early childhood | 51 | 17.0 |
| Middle childhood | 37 | 12.3 |
| Adolescence | 11 | 3.7 |
| Gender |  |  |
| Female | 135 | 45.0 |
| Male | 165 | 55.0 |
| **Patient's type** |  |  |
| Indoor | 101 | 33.7 |
| Outdoor | 100 | 33.3 |
| Emergency | 99 | 33.0 |
| **Most frequent diseases\*** |  |  |
| Different Fevers | 66 | 22.0 |
| Pneumonia | 51 | 17.0 |
| Cough | 48 | 16.0 |
| Delayed Crying | 42 | 14.0 |
| Cold | 40 | 13.3 |
| Vomit | 18 | 6.0 |
| Diarrhea | 16 | 5.3 |
| Neonatal Septicemia | 12 | 4.0 |
| Nasal Congestion | 10 | 3.3 |
| Meningitis | 10 | 3.3 |
| Allergic Problem | 8 | 2.7 |
| Stool Problem | 6 | 2.0 |

\*Several individuals had multiple diseases

### Frequency analysis of prescribed antibiotics

Table 2 below contains frequency and percentage of different antibiotics prescribed by the physicians to the 300 indoor, outdoor and emergency patients who participated in this study. Ceftazidime (14.9%), Amikacin (10.9%), Ceftriaxone (10.4%), Cefixime (9.9%) and Azithromycin (9.4%) were some mostly prescribed antibiotics.

**Table 2.** Frequency and percentage of different Antibiotics

| Antibiotic Name | Frequency | Percentage |
| --- | --- | --- |
| Ceftazidime | 60 | 14.9 |
| Amikacin | 44 | 10.9 |
| Ceftriaxone | 42 | 10.4 |
| Cefixime | 40 | 9.9 |
| Azithromycin | 38 | 9.4 |
| Ampicillin | 33 | 8.2 |
| Gentamicin | 22 | 5.5 |
| Meropenem | 21 | 5.2 |
| Amoxicillin | 14 | 3.5 |
| Cefuroxime | 14 | 3.5 |
| Metronidazole | 14 | 3.5 |
| Flucloxacillin | 13 | 3.2 |
| Erythromycin | 11 | 2.7 |
| Penicillin | 8 | 2.0 |
| Cefaclor | 6 | 1.5 |
| Cefradine | 6 | 1.5 |
| Ciprofloxacin | 6 | 1.5 |
| Clarithromycin | 4 | 1.0 |
| Levofloxacin | 2 | 0.5 |
| Phenoxymethylpenicillin | 2 | 0.5 |
| Cefuroxime + Clavulanic acid | 1 | 0.2 |

### Association between level of doses of antibiotics and patient’s age category, patient type, doses form and, regimen of doses

To check whether prescribed level of doses of antibiotics had any association with patient’s age, patient type, form of doses and regimen of doses, Pearson Chi square test was employed with level of significance 0.05. Table 3 contains the cross-tabulated frequencies and percentages between level of doses of antibiotics and patient’s age, patient type, form of doses and regimen of doses. The Pearson Chi-square test indicated significant statistical association between doses level of antibiotic and patient type with test statistic 62.435 (*df* = 4, *P*<.001), dosage form with test statistic 39.069 (*df* = 4, *P*<.001), regimen of doses with test statistic 18.822 (*df* = 4, *P* = .001) and, age category with test statistic 56.989 (*df* = 10, *P*<.001) (See <u>S1 Table</u>).

**Table 3.** Cross tabulation of level of doses of antibiotics and patient’s age category, patient type, dosage form and, regimen of doses

|  | Level of Doses |  |  |  |  |  |
| --- | --- | --- | --- | --- | --- | --- |
|  | Normal |  | Underdose |  | Overdose |  |
|  | Frequency | Percentage | Frequency | Percentage | Frequency | Percentage |
| **Patient type** |  |  |  |  |  |  |
| Indoor | 70 | 69.3 | 13 | 12.9 | 18 | 17.8 |
| Outdoor | 15 | 15.0 | 30 | 30.0 | 55 | 55.0 |
| Emergency | 50 | 50.5 | 15 | 15.2 | 34 | 34.3 |
| **Dosage form** |  |  |  |  |  |  |
| Solid | 10 | 58.8 | 3 | 17.6 | 4 | 23.5 |
| Solution | 101 | 58.7 | 25 | 14.5 | 46 | 26.7 |
| Suspension | 24 | 21.6 | 30 | 27.0 | 57 | 51.4 |
| **Dosage regimen** |  |  |  |  |  |  |
| 3 or less days | 46 | 64.8 | 6 | 8.5 | 19 | 26.8 |
| 4 to 6 days | 33 | 43.4 | 20 | 26.3 | 23 | 30.3 |
| 7 or more days | 56 | 37.1 | 31 | 20.5 | 64 | 42.4 |
| **Age category** |  |  |  |  |  |  |
| Term Neonatal | 71 | 71.0 | 14 | 14.0 | 15 | 15.0 |
| Infancy | 26 | 38.8 | 15 | 22.4 | 26 | 38.8 |
| Toddler | 10 | 29.4 | 10 | 29.4 | 14 | 41.2 |
| Early childhood | 9 | 17.6 | 8 | 15.7 | 34 | 66.7 |
| Middle childhood | 14 | 37.8 | 9 | 24.3 | 14 | 37.8 |
| Adolescence | 5 | 45.5 | 2 | 18.2 | 4 | 36.4 |

From the cross tabulation, it is evident that for some categories, under dose or overdose of an antibiotic had been prescribed more than normal level of dose. In case of outdoor patients, only 15% had been prescribed for the antibiotics with normal dose level whereas 30% were prescribed under dose and 55% were prescribed overdose, in case of antibiotics that are suspension, only 21.6% had been prescribed for the antibiotics with normal dose level whereas 27.0% were prescribed under dose and 51.4% were prescribed overdose, in case of an antibiotic having regimen of 7 days or more, only 37.1% had been prescribed for the antibiotics with normal dose level but 42.4% were prescribed overdose. Table 3 also shows that normal dose levels and overdose levels had equal percentages for infant patients and patients at middle childhood. Again, 29.4% of the dosage had been prescribed for the antibiotics with normal dose level for the patients at toddler age but 29.4% were prescribed under dose and 41.2% were prescribed overdose, for the patients in early childhood, 17.6% of the dosage had been prescribed for the antibiotics with the normal dose level while 66.7% of the dosage had been prescribed for the antibiotics with overdose.

### Association between frequency of doses of antibiotics and patient’s age category and, patient type

Pearson Chi square test was performed at 0.05 level of significance to find if the prescribed frequency of daily antibiotic consumption had any significant association with patient’s age, patient type, form of doses and regimen of doses. The test suggested that only patient type and patient’s age category had significant association with frequency of antibiotic doses with patient type having test statistic of 20.692 (*df* = 4, *P*<0.001) and patient’s age category having test statistic of 21.868 (*df* = 10, *P* = 0.016) (See <u>S2 Table</u>).

Table 4 contains the cross-tabulated frequencies and percentages between frequency of doses of antibiotics and patient’s age and, patient type. Tabulated data indicates that emergency patients had higher percentage (85.9%) than outdoor patients (58.0%) in case of standard level frequency of antibiotic doses daily consumption. By age category, 65% of the dosage have been prescribed for the antibiotics with standard frequency for the patients at term neonatal, 59.7% for the patients at infancy, 70.6% for the patients at toddler age, 84.3% for the patients at early childhood, 81.1% for the patients at middle childhood and 72.7% of the dosage have been prescribed for the antibiotics with standard frequency for the patients at adolescence. Although not much of the antibiotics have been prescribed to consume more frequently than medically suggested frequency, but less frequency of consumption than suggested had been prescribed at high percentage.

**Table 4.** Cross tabulation of frequency of doses of antibiotics and patient’s age category and, patient type

|  | Frequency of Doses |  |  |  |  |  |
| --- | --- | --- | --- | --- | --- | --- |
|  | Less frequent |  | Standard |  | More frequent |  |
|  | Frequency | Percentage | Frequency | Percentage | Frequency | Percentage |
| **Patient type** |  |  |  |  |  |  |
| Indoor | 30 | 29.7 | 67 | 66.3 | 4 | 4.0 |
| Outdoor | 35 | 35.0 | 58 | 58.0 | 7 | 7.0 |
| Emergency | 10 | 10.1 | 85 | 85.9 | 4 | 4.0 |
| **Age category** |  |  |  |  |  |  |
| Term Neonatal | 32 | 32.0 | 65 | 65.0 | 3 | 3.0 |
| Infancy | 18 | 26.9 | 40 | 59.7 | 9 | 13.4 |
| Toddler | 9 | 26.5 | 24 | 70.6 | 1 | 2.9 |
| Early childhood | 7 | 13.7 | 43 | 84.3 | 1 | 2.0 |
| Middle childhood | 6 | 16.2 | 30 | 81.1 | 1 | 2.7 |
| Adolescence | 3 | 27.3 | 8 | 72.7 | 0 | 0.0 |

### Deviation of dose from standard dose amount by Antibiotics

The following Table 5 shows the median deviation of dose from their standard level for every antibiotic indicating which antibiotic, when prescribed, was much deviated from the standard level suggested. Some antibiotics were prescribed with high median deviation from their suggested level, e.g., Flucloxacillin had 100% median deviation from the standard level and Cefixime’s median dose deviation was 50.95% from the standard level. On the contrary, Ampicillin, Ceftazidime, Ceftriaxone, Meropenem, and some other doses have 0% median dosing deviation from normal.

**Table 5.** Median deviation of antibiotic’s dose from standard dose level

| Antibiotic Name | Median Deviation |
| --- | --- |
| Flucloxacillin | 100.00% |
| Cefixime | 50.95% |
| Erythromycin | 41.18% |
| Levofloxacin | 37.50% |
| Azithromycin | 25.00% |
| Gentamicin | 20.00% |
| Ciprofloxacin | 15.47% |
| Amoxicillin | 12.08% |
| Cefuroxime | 11.50% |
| Phenoxymethylpenicillin | 11.11% |
| Clarithromycin | 9.92% |
| Metronidazole | 8.84% |
| Amikacin | 3.97% |
| Cefaclor | 3.27% |
| Ampicillin | 0.00% |
| Ceftazidime | 0.00% |
| Ceftriaxone | 0.00% |
| Cefradine | 0.00% |
| Meropenem | 0.00% |
| Penicillin | 0.00% |

**Figure 1.**
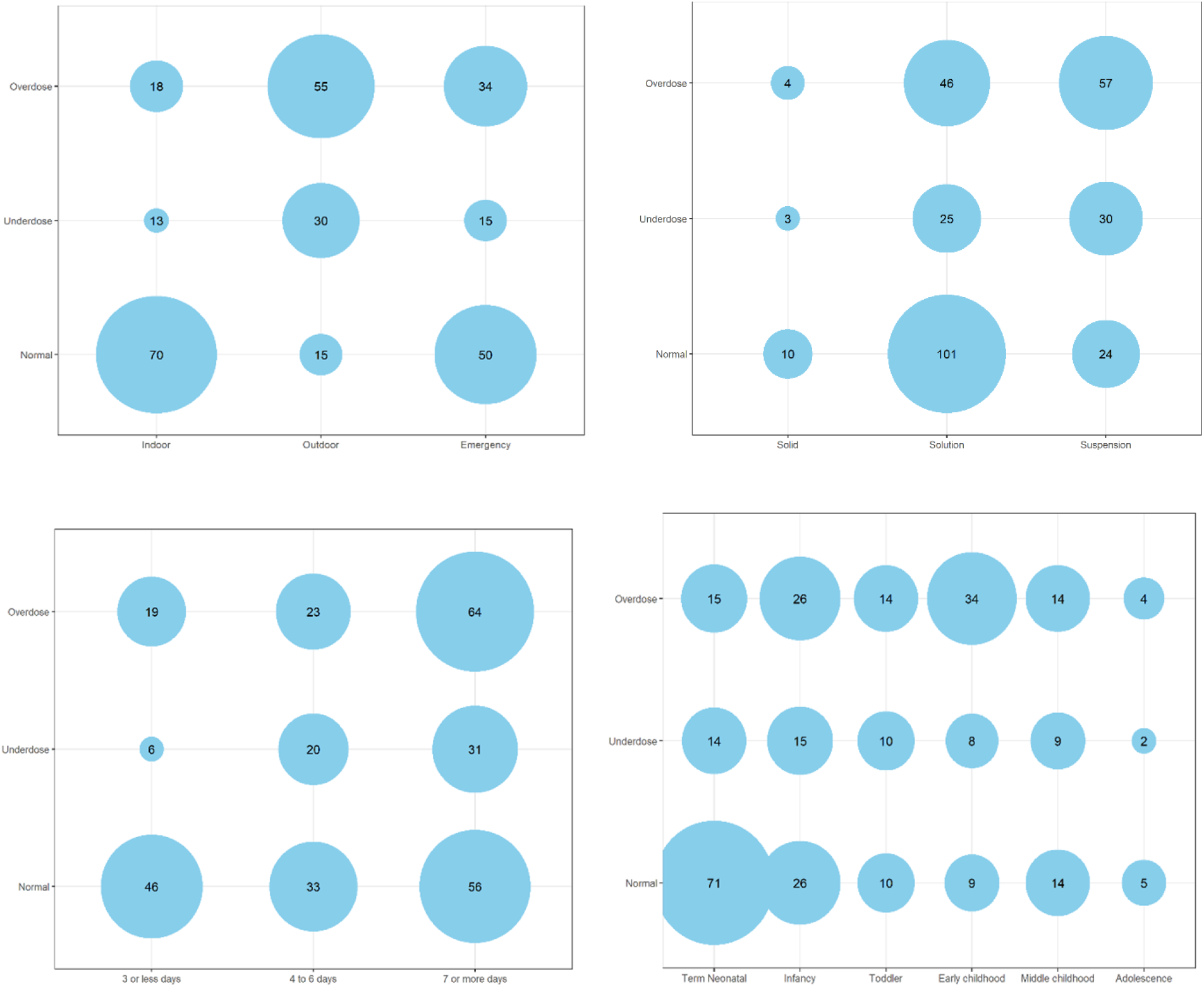
Visualization of cross tabulation between level of doses of antibiotics and patient’s age category, patient type, dosage form and, regimen of doses

**Figure 2.**
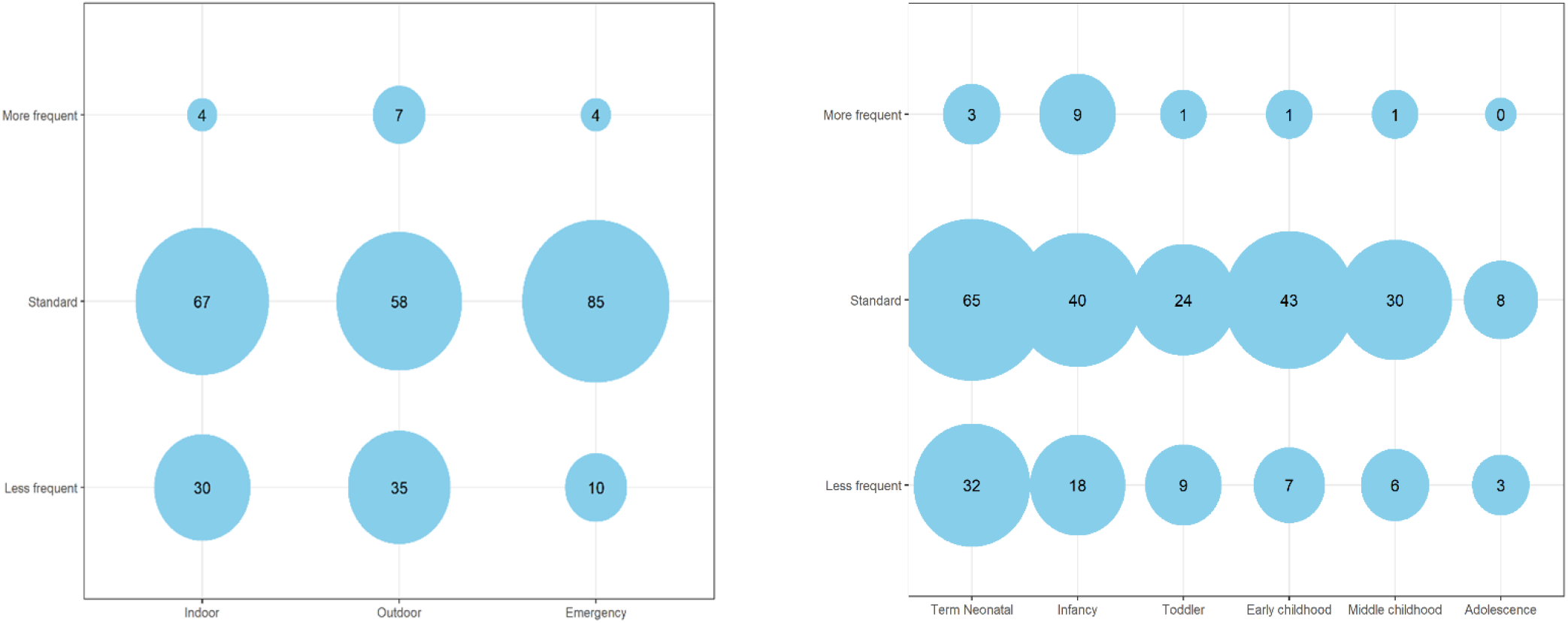
Visualization of cross tabulation between frequency of doses of antibiotics and patient’s age category and, patient type

### Test of deviation of dose from the standard dose amount by patient type and age

The deviation of the dose amount from the standard amount has been shown by the median absolute deviation of dose amount from the standard. Deviation of dose level were also checked by patient type and age, to see if there were any significant difference in median absolute deviation of antibiotic dose by patient type and their age category. It was found that indoor patients have a 0% median absolute deviation and outdoor patients had 25% median absolute dosing deviation, which was much higher than indoor patients. But, for emergency patients, deviation was 31.19%, which is higher in number than outdoor patients and indoor patients. The Kruskal-Wallis test, at 0.05 level of significance, indicates that the median absolute deviations were significantly different by patient type with test statistic 62.55 (df = 2, *P*<0.001). Outdoor patients and emergency patients had a much higher deviation in dose amount than standard level of antibiotic’s dose compared to indoor patients.

In case of the median absolute deviation of dose by age category, it was found that the term neonatal patient’s median absolute dosing deviation was 0%. For toddlers, the median absolute dosing deviation was 7.63%, and for middle childhood, 11.11%. But for the patients in early childhood, the median absolute deviation of dose from the standard was 33.93%, for infants, 37.08%, and for adolescents, 50%. So, patients in early childhood, infancy, and adolescence had higher dosing median absolute deviation from standard amount than the patients at term neonatal and toddler. The Kruskal-Wallis test, at 0.05 level of significance, with test statistic 62.721 (df = 5, *P*<0.001) then indicates that the median absolute deviations were significantly different for different age groups of the patients (See <u>S3 Table</u>).

## Discussion

A heterogeneous pediatric population underscores the importance of prescribing multiple dosage forms. Cover up the wide range of pediatric populations; prescribers prescribed solution, suspension, and solid dosage forms of different antibiotics. During the prescribing of various antibiotics for different pediatrics categories, most prescribers preferred liquid preparations over solid types. The research findings revealed that inappropriate antibiotic doses’ incidences happened with solution and suspension types of antibiotic dosage forms in pediatric patients. Previous studies described those liquid preparations were preferable because of their maximal dosing flexibility, and these can easily use over a wide range of ages, including neonates (13).

This research showed a ranking of the top 12 diagnoses of diseases that commonly occurred among pediatric patients of different ages. Among these top 12 diseases for which various antibiotics were prescribed, fever ranked first and allergic problem and stool problem simultaneously ranked last. All these diseases were common among all categories of pediatric patients. While dealing with these diseases using commercially available antibiotic dosages, prescribers prescribed a higher level of antibiotic doses for patients at the outdoor unit, followed by patients at the emergency unit and patients at the indoor unit. On the other hand, prescribed antibiotics with under doses were higher among the outdoor pediatric patients. However, prescribed normal doses of antibiotics were higher among the indoor unit patients and lowered among the outdoor unit patients.

This research’s experimental data lucidly indicated that antibiotics with high doses were recommended for durations of ≥7days and followed by 4 to 6 days. This research also showed that prescribed antibiotics of under doses were recommended for durations of 4 to 6 days, followed by ≥7days and ≤3 days, respectively. This study clearly showed a trend of decreasing the average level of antibiotic doses with the increasing dosage regimen.

Early childhood patients had a higher level of antibiotic doses in comparison with other age groups. Patients at toddler age had the highest percentage of under level of any antibiotic doses. According to this finding, it can easily be interpreted that prescriber miscalculated the doses and were unable to adjust the right amount of dose for the patients at early childhood and toddler age while manipulating the commercially available doses of antibiotics. Prescribers who were unfamiliar with the proper pediatric guidelines could have a chance to commit such medication dosing errors over and over again.

Appropriate or correct doses of antibiotics can ensure safety and successful therapeutic outcomes in pediatric patients. This research observed that three antibiotics named cefuroxime plus clavulanic acid, cephradine, and penicillin were prescribed at normal doses per a prescription order. There were 6 more antibiotics, half or more of which were named ampicillin, cefaclor, ceftazidime, ceftriaxone, levofloxacin, meropenem, etc. were prescribed at normal doses. Out of a total of 21 different types of prescribed antibiotics for pediatric patients, observation showed that prescribers unwittingly gave every dose of phenoxymethyl penicillin at a dose higher than the recommended dose. Another type of beta-lactam 3rd generation cephalosporin named cefixime, 80 percent of which was prescribed at higher dose for pediatric patients. There were other certain types of prescribed antibiotics, half or more of which such as ciprofloxacin, flucloxacillin, gentamicin, azithromycin, etc. were prescribed at higher doses level. However, other certain types of prescribed antibiotics, half or more of which such as amikacin, cefuroxime, clarithromycin, levofloxacin, metronidazole, etc. were prescribed at under doses than the recommended doses. The higher administering percentages of inappropriate doses of antibiotics may affect the health of the pediatric population by enabling the development of antibiotic resistance or therapeutic failures or certain antibiotic-induced adverse effects. A renowned daily newspaper in Bangladesh named The Daily Star on January 13 in 2020 published an article headlining antibiotics use, sale: who needs prescription? where they investigated the inappropriate and overuse of antibiotics and found that ceftriaxone, a watch group drug, used most frequently by people in Bangladesh (21). This research work also reported the most used 3^rd^generation cephalosporin named ceftazidime and ceftriaxone among pediatric patients. Flucloxacillin is known for inducing hepatotoxicity among pediatric patients (22). Scott SI and his colleagues in 2016 observed that the repeating exposure of different antibiotics in the first 2 years of life of a child is associated with an increase in the risk gaining obesity (23). Unnecessary antibiotics and antibiotics at inappropriate doses can also trigger hazardous consequences among pediatric patients.

Among all the prescribed antibiotics, flucloxacillin has the highest or all the prescribed flucloxacillin for pediatric patients has shown median deviation from the standard followed by cefixime, erythromycin, levofloxacin, azithromycin, gentamycin, ciprofloxacin, amoxicillin, cefuroxime, phenoxymethylpenicillin, clarithromycin, metronidazole, amikacin, and cefaclor, etc. all had clearly demonstrated median dosing deviation from normal. However, there were other certain prescribed antibiotics for pediatric patients like ampicillin, ceftazidime, ceftriaxone, meropenem, penicillin, etc. have no median deviation from the standard dose levels.

Standard frequencies for different antibiotics had been reported with a high rate among patients in early childhood and in emergency care unit. Maintenance of standard frequencies for different antibiotics was reported lower among patients at infancy and in outdoor care unit. Generally, a child’s waking day is as little as 12h which is considered much shorter than that of an adult (22). So pediatric patients’ sleeping hours will be affected if they are recommended more frequent antibiotic doses. That is why assessing the most appropriate interval is considered one of the prime factors to be needed to evaluate when selecting a drug dosage regimen for a pediatric patient (22).

Inappropriate antibiotic doses may pose a threat to pediatric patients in the emergency care unit. In this modern medical health system facility, improper dosing for the pediatric patient population is one of the most common medication errors (24, 25). Inconsistencies in growth rates among pediatric patients of different ages also may act as a prime factor for arising confusion among prescribers to quickly recognize when a dose of antibiotic is incorrect or inappropriate even in patients of a similar age. In emergency patient care, limited available patient information, disease complexity, higher patient turnover, lack of knowledge of appropriate pediatric drug references, etc., can act as significant barriers to selecting proper doses of antibiotics for pediatric patients. Without knowing pediatric patients’ weight, prescribers cannot select or properly evaluate any appropriate antibiotic doses. This study revealed that the chosen deviation of different antibiotics’ doses amounts was higher among the emergency department’s pediatric patients. In this research, patients in early childhood, infancy, and adolescence have higher dosing median deviation from standard amount than the patients at term neonatal and toddler.

This study only aimed to investigate the use of antibiotics dose levels among pediatric patients. We can accomplish some other further studies like consequences or possible adverse effects of these antibiotics among pediatric patients. Future endeavors can be to investigate whether the prescribed higher doses of the identified antibiotics induce any serious adverse effects among pediatric patients in Bangladesh. Another future endeavor will be to explore the hazardous impacts on pediatric patients’ health and development from the consumption of several unnecessary antibiotics at inappropriate doses.

## Conclusion

The occurrence of various medication errors is a common occurrence among pediatric patients. Miscalculations and neglect in following proper pediatric guidelines when prescribing antibiotics for pediatric patients may contribute to these errors. Therefore, maintaining an adequately calculated weight range based on pediatric antibiotic standard dosing practice in all clinical settings for treating a heterogeneous pediatric population may reduce the likelihood of such instances of inappropriate dosing among pediatric patients. If prescribers do not strictly adhere to the pediatric dosing guidelines when prescribing antibiotics, adverse drug reactions and therapeutic failures will be extremely common among pediatric patients. Age, weight, and body surface area of the pediatric patient are essential factors to consider when selecting a drug dosage regimen or effective doses. A missing or incorrect patient weight can also result in a medication dosage that differs significantly from the desired dose and has harmful health consequences. Therefore, without considering the precise weight of pediatric patients, prescribers cannot choose or measure appropriate antibiotic dosages. To ensure good clinical pharmacy practice in all settings, prescribers should allow parents or caregivers of pediatric patients to be their active partners in minimizing the adverse effects of inappropriate antibiotic doses, when appropriate.

In Bangladesh, the lack of age-appropriate concentrations of formulations and commercially available age-appropriate products may result in inappropriate antibiotic dosages or serious dosing errors. Only precise dose measurement while performing any additional dose calculations and manipulations on commercially available antibiotic doses can reduce the incidence of pediatric patients receiving inappropriate antibiotic doses. This clinical study demonstrated unequivocally that maximizing the contribution of clinical pharmacists and the dissemination of timely, relevant information while supporting the treatment decision-making process of a pediatric patient not only decreases the percentage of prescriptions containing inappropriate antibiotic doses or dose errors but also ensures the existence of good clinical pharmacy practice in pediatric patient care units.

## Data Availability

All data produced in the present study are available upon reasonable request to the authors

## Supporting information

**S1 Table. Significance of association between level of doses of antibiotics and patient’s age category, patient type, dosage form and, regimen of doses**

(DOCX)

**S2 Table. Significance of association between frequency of doses of antibiotics and patient’s age category and, patient type**

(DOCX)

**S3 Table: Test of deviation of dose from the standard dose amount**

(DOCX)

**S1 File. Study questionnaire in Bangla**

(DOCX)

**S2 File. Study questionnaire in English**

(DOCX)

## Acknowledgment

The authors would like to give gratitude to all the respondents. And they would like to express gratefulness to all the individuals who have contributed to this research directly or indirectly.

## Data availability statement

All data are freely accessible without any restriction.

